# Right and Left Bundle Branch Block in the General Population: Structural Heart Findings, Clinical and Prognostic Implications: Insights from the Hamburg City Health Study

**DOI:** 10.64898/2026.09.07.26362475

**Authors:** Juliana Senftinger, Vivian Link, Matthias Klimek, Laura Dorothea Keil, Maria Luisa Benesch Vidal, Jan Rieß, Raphael Twerenbold, Christina Magnussen, Renate B. Schnabel, Stefan Blankenberg, Andreas Ziegler, Peter Moritz Becher, Peter Clemmensen

**Affiliations:** Department of Cardiology, University Heart & Vascular Center Hamburg, University Medical Center Hamburg-Eppendorf, Hamburg, Germany; Center for Population Health Innovation (POINT), University Heart & Vascular Center Hamburg, University Medical Center Hamburg-Eppendorf, Hamburg, Germany; Cardio-CARE, Medicine Campus Davos, Davos, Switzerland; Germany Center for Cardiovascular Research (DZHK), Partner Site North, Hamburg, Germany; Discipline of Statistics, School of Agriculture and Science, University of KwaZulu-Natal, Pietermaritzburg, South Africa; Department of Medicine-Cardiology, Zealand University Hospital, Nykoebing F, Denmark

**Keywords:** bundle branch blocks, general population, echocardiography, prognostic implication

## Abstract

**Background:** This study aimed to determine the prevalence of BBB, characterize associated clinical and echocardiographic phenotypes, and evaluate associations with incident cardiovascular diseases and all-cause mortality in the general population.

**Methods:** We analyzed 12-lead electrocardiograms from 14,212 subjects from the Hamburg City Health Study (45-74 years). BBBs were defined using Minnesota Code criteria. We applied elastic net regularization to identify clinical associations, and multivariable Cox models for outcome analyses. The total cardiovascular outcome included all-cause mortality, incident heart failure, myocardial infarction, atrial fibrillation, pulmonary embolism, and stroke.

**Results:** Complete left (cLBBB) and right (cRBBB) bundle branch blocks were prevalent in 0.8% and 2.3% of subjects, respectively. Subjects with complete BBB were generally older with a higher burden of cardiovascular risk factors and comorbidities, cLBBB additionally showed higher N-terminal prohormone of brain natriuretic peptide levels.

The main predictor of BBB was age. Male sex was an additional predictor for cRBBB, while hypertension, hypercholesterolemia, and reduced left ventricular ejection fraction predicted cLBBB. Both, cLBBB and cRBBB were associated with a reduced left ventricular ejection fraction, increased septal wall thickness, and left atrial enlargement. cRBBB demonstrated right ventricular dilation, whereas cLBBB showed markedly impaired left atrial strain.

In adjusted analyses, both BBBs were associated with the total cardiovascular outcome (cLBBB: hazard ratio [HR], 2.71; 95% confidence interval [CI], 1.68–4.39; p<0.001; cRBBB: HR, 1.52; 95% CI, 1.09–2.12; p=0.01). Although both were significantly associated with incident heart failure, myocardial infarction, and atrial fibrillation, cLBBB uniquely was associated with all-cause mortality, whereas cRBBB distinctly was associated with incident stroke.

**Conclusion:** In the general population, BBB is a high-risk phenotype characterized by subclinical cardiac alterations and a markedly worse long-term outcome. These findings underscore the importance of early risk stratification. Proactive clinical management in otherwise asymptomatic individuals presenting with these conduction disturbances should be considered.

## 1. Introduction

Bundle branch blocks (BBBs) represent a disorder of electrical conduction within the His-Purkinje system, resulting in delayed ventricular depolarization.^1^ Although the overall prevalence of right and left bundle branch blocks (RBBB and LBBB) in the general population is relatively low, their incidence increases significantly with advancing age and it is higher in men.^2–4^ In cohorts with established cardiovascular diseases, the presence of a BBB is well-documented and consistently associated with adverse cardiovascular outcomes.^5–7^

Evidence on the prognostic implications of BBB in the general population remains remarkably limited. For LBBB, a small number of predominantly older studies suggest an association with adverse outcomes, particularly an increased incidence of heart failure (HF).^8^ Yet, the differential diagnosis of a purely idiopathic BBB remains challenging, complicating definitive prognostic stratification.^4^ Conversely, RBBB has historically been classified as a benign electrocardiographic variant with recent investigations challenging this paradigm, indicating a potential association with unfavorable long-term outcomes, including higher rates of subsequent pacemaker implantation.^9^

The causality and temporal sequence of their pathogenesis often remain unsolved. A central clinical question whether intraventricular conduction delay itself induces secondary structural remodeling, or whether the BBB is an early electrocardiographic correlate of a primary yet subclinical structural heart disease, including subclinical _HF._10,11

Therefore, the aim of the present study is to comprehensively characterize BBBs in the general middle-aged population in terms of clinical implications, underlying structural changes determined by 2-dimensional (2D) echocardiography, and their prognostic implications.

## 2. Methods

### 2.1 Study Setting and Population

Data from the first 16,411 subjects enrolled in the Hamburg City Health Study (HCHS; www.hchs.hamburg) were used for this analysis. The HCHS (ClinicalTrials.gov identifier: NCT03934957) is an ongoing, prospective, single-center, population-based cohort study conducted in Hamburg, Germany, featuring random subject selection and long-term follow-up. The study investigates interactions among socioeconomic risk factors, advanced imaging modalities, physiological measurements, and clinical variables. A detailed description of the study design has been published previously.^12^

For the present analysis, after excluding individuals with missing electrocardiographic (ECG) data, rhythms other than sinus rhythm, or pacemaker stimulation, the final study population comprised 14,212 subjects (**Figure S1**). Baseline covariates included demographic data, comorbidities, and biomarkers (e.g., N-terminal prohormone of brain natriuretic peptide [NT-proBNP], high-sensitivity C-reactive protein [hsCRP], high-sensitivity Troponin I [hsTnI], estimated Glomerular Filtration Rate [eGFR]). Medications were classified using Anatomical Therapeutic Chemical (ATC) codes and included antihypertensive, HF, antiarrhythmic, and statin therapies. The ethics committee of the State of Hamburg (Chamber of Medical Practitioners, reference number PV5131) did not object against the conduct of the study, and the respective Data Protection Commissioners agreed to the participation of the study. All subjects provided written informed consent.

### 2.2 ECG and Transthoracic Echocardiography (TTE)

The cardiovascular assessments included a digital 12-lead ECG (with a 2-minute rhythm strip) and baseline TTE. Anonymized ECGs were manually evaluated by a physician for standard parameters, pathological findings were measured a second time using an online tool (Schiller AG, https://cloud.schiller.ch/apps/user). BBB were defined according to the Minnesota Code criteria.^13,14^

TTE was performed by certified professionals using a Siemens Acuson SC2000 Prime device in accordance with the American Society of Echocardiography (ASE) and the European Association of Cardiovascular Imaging (ASE/EACVI) guidelines. All TTE data were quality-controlled using literature-based clinical cut-offs and dependencies. All inconsistencies were reassessed by medical professionals to establish a final quality-controlled data set.^14^

### 2.3 Follow-up and Clinical Outcomes

Between October 2022 and April 2023, a comprehensive follow-up assessment was conducted to determine targeted clinical endpoints (morbidity) and vital status (mortality). This follow-up included subjects who had completed their baseline HCHS examination prior to January 1, 2022.

Vital status was ascertained through population registries and reports from relatives. In the event of death, the cause was identified via the public health department and cross-referenced with the Department of Forensic Medicine at the University Medical Center Hamburg-Eppendorf. For morbidity follow-up, subjects completed questionnaires assessing selected incident diseases and their corresponding dates of diagnosis. Self-reported endpoints were subsequently adjudicated using multiple data sources, including hospital information systems, medical discharge letters, and direct inquiries to the subjects’ general practitioners and specialists. Median follow-up was 4.3 years (95% confidence interval [CI], 3.0–5.8) for subjects without BBB, 4.7 years (95% CI, 3.7–6.1) for those with complete LBBB (cLBBB), and 4.5 years (95% CI, 3.6–6.0) for those with complete (cRBBB).

The total cardiovascular outcome was defined as a composite of all-cause mortality, incident myocardial infarction (MI), stroke, HF, atrial fibrillation (AF), or pulmonary embolism. The secondary focused cardiovascular outcome was a composite of cardiovascular mortality, MI, or stroke. All-cause mortality was additionally evaluated as an independent outcome.

### 2.4 Statistical Analysis

Baseline characteristics, stratified by BBB, are presented as medians with quartiles in parenthesis for continuous variables, and counts with percentages for categorical variables. Elastic net–penalized regression was used for variable selection in descriptive regression models for cRBBB and cLBBB. This was followed by a standard logistic regression, where p-values were Bonferroni-corrected. Potential non-linear associations between continuous QRS duration and clinical outcomes were evaluated using fractional polynomials within Cox regressions. Time-to-event analyses used attained age as time scale. Cox proportional hazards models were used to evaluate all-cause mortality. For other causes of death and cardiovascular mortality as competing events, Fine-Gray subdistribution hazard models were employed to account for competing risks. For these analyses, sex was included as a covariate and patients with prevalent MI, HF, or stroke were excluded. Cumulative incidence curves were generated. All analyses were performed using R (version 4.6.0, R Foundation for Statistical Computing, Vienna, Austria). The nominal type I error level was 0.05. Further methodological details are provided in the supplementary material.

## 3. Results

### 3.1 Prevalence of BBB

Among 14,212 subjects, incomplete RBBB (iRBBB) was present in 2.1% and cRBBB in 2.3%, whereas incomplete (iLBBB) and complete LBBB were observed in 0.1% and 0.8%, respectively. Except for iRBBB, subjects with BBB were older, and except from cLBBB, all subtypes showed a male predominance (**Figure 1**).

**Figure 1:**
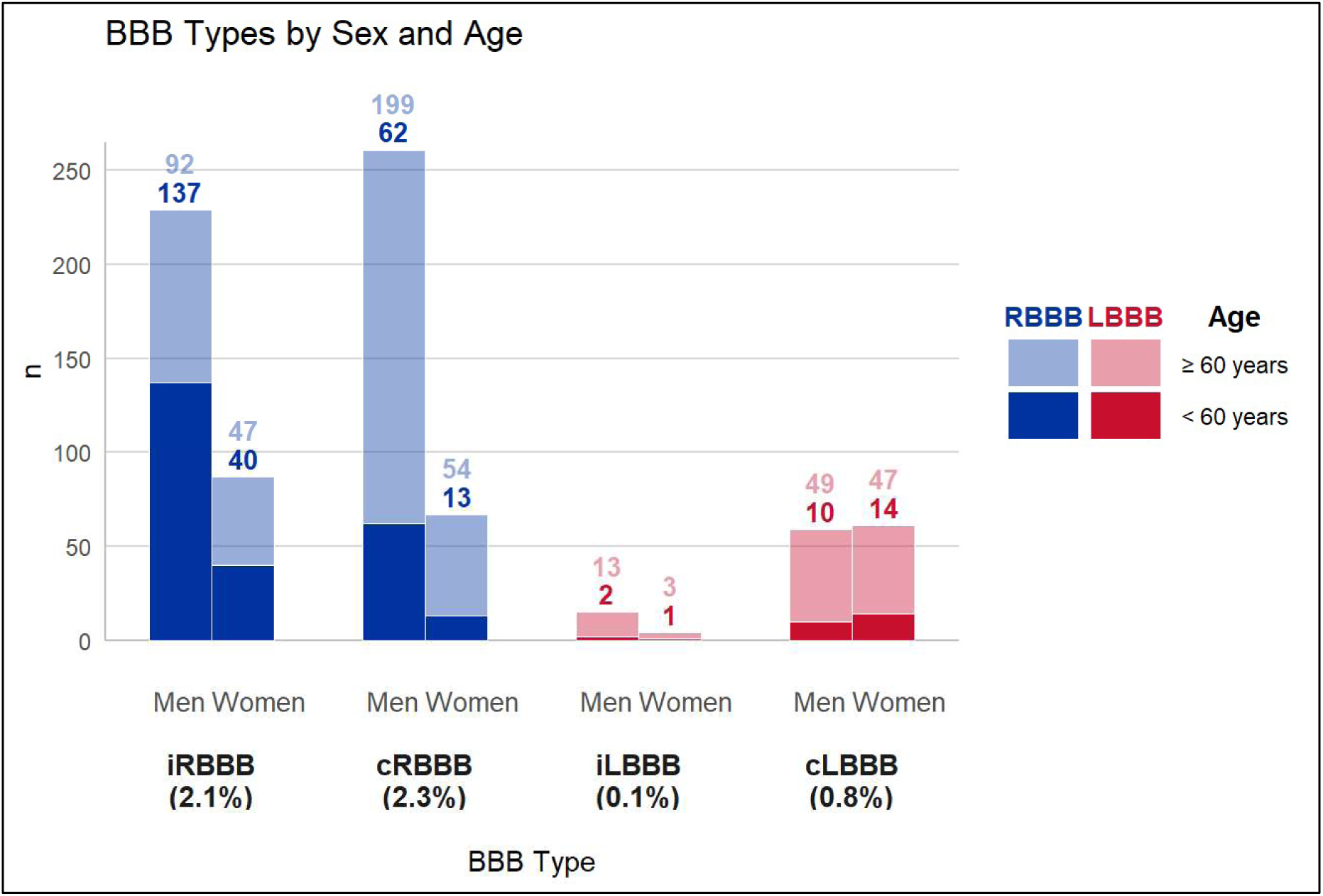
Age and Sex Distribution of Bundle Branch Block Subtypes in the General Population. Bar graph illustrating the prevalence of complete and incomplete RBBB (blue) and LBBB (red) within the total cohort (n = 14212) according to sex and age. *Abbreviations: cLBBB = complete left bundle branch block; cRBBB = complete right bundle branch block; iLBBB = incomplete left bundle branch block; iRBBB = incomplete right bundle branch block*.

### 3.2 Patient Characteristics

Individuals with cRBBB, iLBBB, and cLBBB exhibited a greater burden of comorbidities and a higher prevalence of cardiovascular risk factors, including hypertension, hypercholesterolemia, and diabetes mellitus, whereas the iRBBB group demonstrated cardiovascular risk profiles comparable to those of the total cohort (**Table 1**). Compared with individuals without conduction defects, complete BBB were associated with a higher prevalence of known coronary artery disease (no BBB 3.9%, cRBBB 12.2%, cLBBB 11.4%), and isolated cLBBB was additionally associated with a higher prevalence of a history of HF accompanied by higher baseline NT-proBNP levels compared to individuals without BBB (HF: no BBB 2.2%, cLBBB 17.6%; NT-proBNP: No BBB 73.0 [quartiles 42.0-131.0] ng/l; cLBBB 171.0 [quartiles 67.5-260.0] ng/l) (**Table 1 and Table S1**).

**Table 1:**
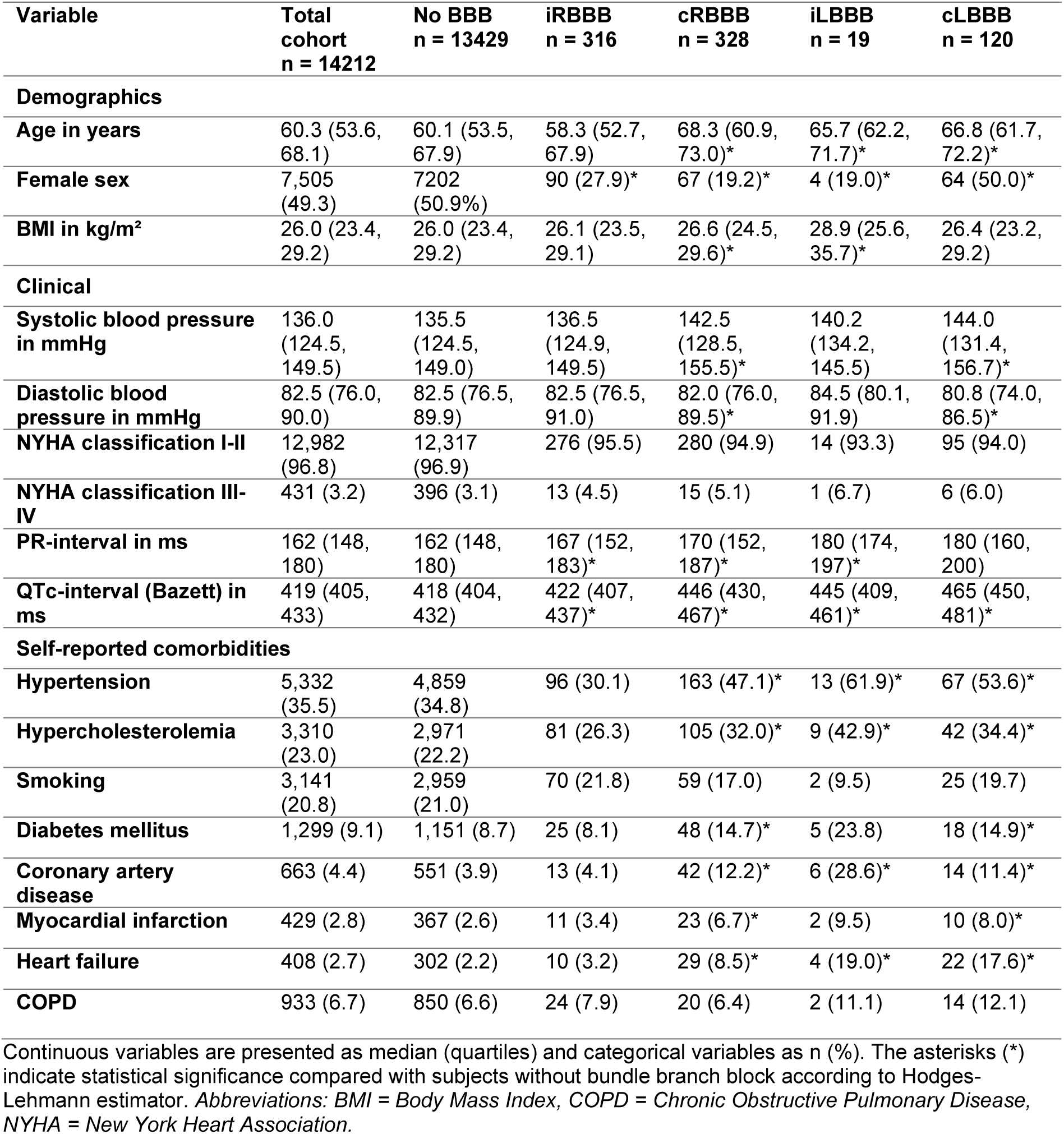
Baseline Characteristics by Bundle Branch Block Type.

| Variable | Total cohort<br>n = 14212 | No BBB<br>n = 13429 | iRBBB<br>n = 316 | cRBBB<br>n = 328 | iLBBB<br>n = 19 | cLBBB<br>n = 120 |
| --- | --- | --- | --- | --- | --- | --- |
| **Demographics** |  |  |  |  |  |  |
| **Age in years** | 60.3 (53.6, 68.1) | 60.1 (53.5, 67.9) | 58.3 (52.7, 67.9) | 68.3 (60.9, 73.0)* | 65.7 (62.2, 71.7)* | 66.8 (61.7, 72.2)* |
| **Female sex** | 7,505 (49.3) | 7202 (50.9%) | 90 (27.9)* | 67 (19.2)* | 4 (19.0)* | 64 (50.0)* |
| **BMI in kg/m<sup>2</sup>** | 26.0 (23.4, 29.2) | 26.0 (23.4, 29.2) | 26.1 (23.5, 29.1) | 26.6 (24.5, 29.6)* | 28.9 (25.6, 35.7)* | 26.4 (23.2, 29.2) |
| **Clinical** |  |  |  |  |  |  |
| **Systolic blood pressure in mmHg** | 136.0 (124.5, 149.5) | 135.5 (124.5, 149.0) | 136.5 (124.9, 149.5) | 142.5 (128.5, 155.5)* | 140.2 (134.2, 145.5) | 144.0 (131.4, 156.7)* |
| **Diastolic blood pressure in mmHg** | 82.5 (76.0, 90.0) | 82.5 (76.5, 89.9) | 82.5 (76.5, 91.0) | 82.0 (76.0, 89.5)* | 84.5 (80.1, 91.9) | 80.8 (74.0, 86.5)* |
| **NYHA classification I-II** | 12,982 (96.8) | 12,317 (96.9) | 276 (95.5) | 280 (94.9) | 14 (93.3) | 95 (94.0) |
| **NYHA classification III-IV** | 431 (3.2) | 396 (3.1) | 13 (4.5) | 15 (5.1) | 1 (6.7) | 6 (6.0) |
| **PR-interval in ms** | 162 (148, 180) | 162 (148, 180) | 167 (152, 183)* | 170 (152, 187)* | 180 (174, 197)* | 180 (160, 200) |
| **QTc-interval (Bazett) in ms** | 419 (405, 433) | 418 (404, 432) | 422 (407, 437)* | 446 (430, 467)* | 445 (409, 461)* | 465 (450, 481)* |
| **Self-reported comorbidities** |  |  |  |  |  |  |
| **Hypertension** | 5,332 (35.5) | 4,859 (34.8) | 96 (30.1) | 163 (47.1)* | 13 (61.9)* | 67 (53.6)* |
| **Hypercholesterolemia** | 3,310 (23.0) | 2,971 (22.2) | 81 (26.3) | 105 (32.0)* | 9 (42.9)* | 42 (34.4)* |
| **Smoking** | 3,141 (20.8) | 2,959 (21.0) | 70 (21.8) | 59 (17.0) | 2 (9.5) | 25 (19.7) |
| **Diabetes mellitus** | 1,299 (9.1) | 1,151 (8.7) | 25 (8.1) | 48 (14.7)* | 5 (23.8) | 18 (14.9)* |
| **Coronary artery disease** | 663 (4.4) | 551 (3.9) | 13 (4.1) | 42 (12.2)* | 6 (28.6)* | 14 (11.4)* |
| **Myocardial infarction** | 429 (2.8) | 367 (2.6) | 11 (3.4) | 23 (6.7)* | 2 (9.5) | 10 (8.0)* |
| **Heart failure** | 408 (2.7) | 302 (2.2) | 10 (3.2) | 29 (8.5)* | 4 (19.0)* | 22 (17.6)* |
| **COPD** | 933 (6.7) | 850 (6.6) | 24 (7.9) | 20 (6.4) | 2 (11.1) | 14 (12.1) |

Continuous variables are presented as median (quartiles) and categorical variables as n (%). The asterisks (\*) indicate statistical significance compared with subjects without bundle branch block according to Hodges-Lehmann estimator. *Abbreviations: BMI = Body Mass Index, COPD = Chronic Obstructive Pulmonary Disease, NYHA = New York Heart Association.*

### 3.3 Factors Associated with Prevalent cRBBB and cLBBB

We evaluated self-reported risk factors, including age, sex, hypertension, hypercholesterolemia, diabetes mellitus, and left ventricular ejection fraction (LVEF). For cRBBB, higher age and male sex were identified as significant covariates, yielding a discrimination model with an area under the receiver operating characteristic curve (AUC) of 0.75 (95% CI 0.72-0.78).

For cLBBB, all evaluated risk factors except sex and diabetes mellitus were retained in the elastic net model. A reduced LVEF emerged as the strongest predictive feature. Together, this group of predictive covariates discriminated cLBBB from non-LBBB cases, achieving an AUC of 0.79 (95% CI 0.74-0.84) (**Figure 2**).

**Figure 2:**
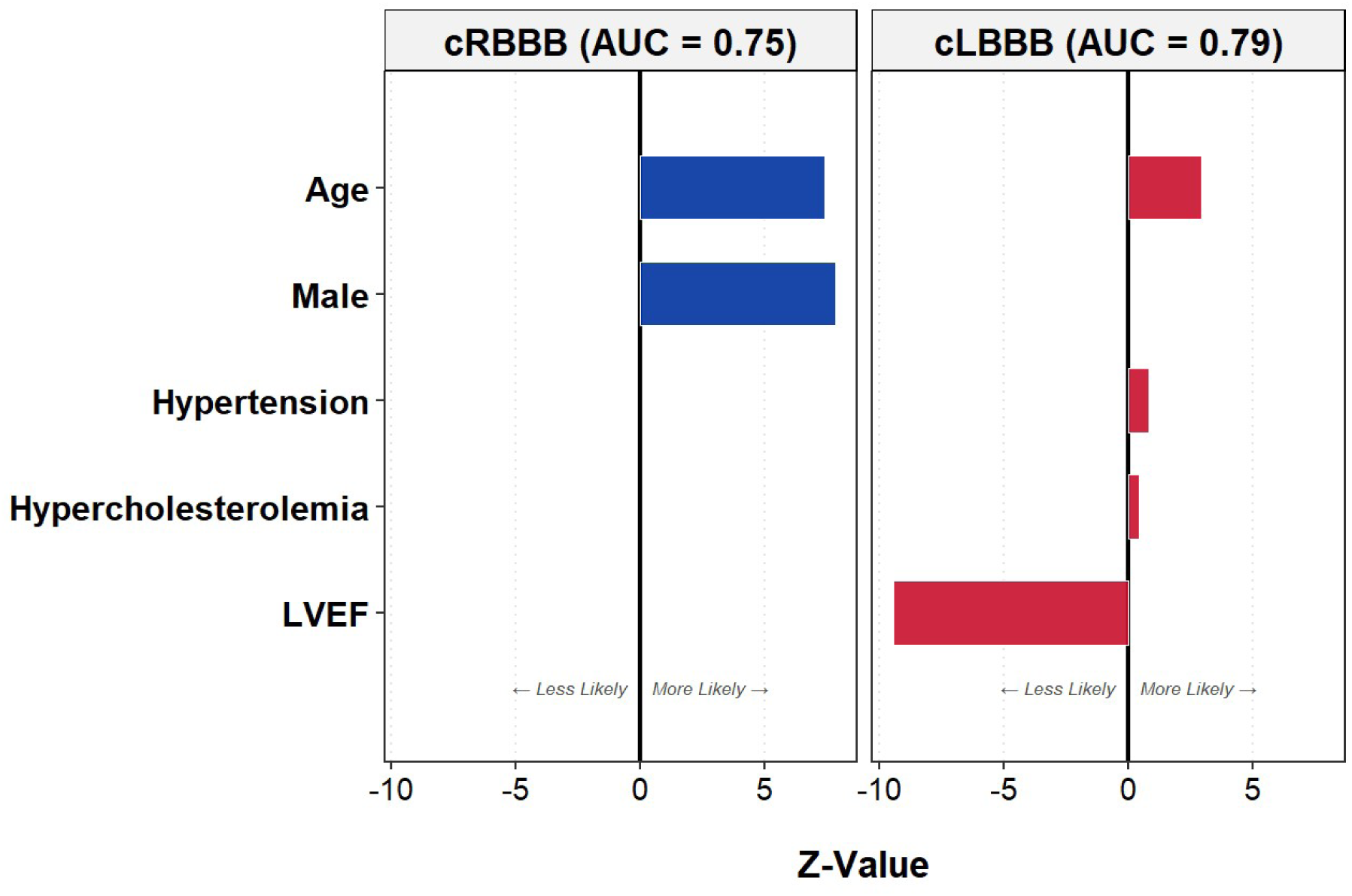
Standardized Effect Sizes of Clinical Predictors for Right and Left Bundle Branch Blocks. Bar plots illustrate the Z-values of clinical variables predicting right bundle branch block (left panel) and left bundle branch block (right panel). The x-axis indicates directionality: negative Z-values represent factors associated with a decreased likelihood of the condition, whereas positive Z-values denote factors associated with an increased likelihood. *Abbreviations: AUC = area under the receiver operating characteristic curve; cLBBB = complete left bundle branch block; cRBBB = complete right bundle branch block; LVEF = left ventricular ejection fraction*.

### 3.4 Electrocardiographic and 2D-Echocardiographic Parameters

In the total cohort, QRS duration demonstrated a narrow distribution, peaking at 92 milliseconds [ms] (standard deviation [SD] 10.1ms). In subjects with RBBB, the mean QRS duration was 146ms (SD 13.7ms), and individuals with LBBB had even wider QRS complexes with a mean of 146.0ms (SD 15.9ms), reaching maximum values of up to 200ms (**Figure S2**). Compared with the total cohort, PR-intervals (no BBB: 162 [quartiles 148-180]ms, cRBBB 180 [quartiles 152-187] ms, iLBBB: 180 [quartiles 174-197] ms, cLBBB: 180 [quartiles 160-200] ms) and QTc intervals (no BBB: 418 [quartiles 404-432] ms, cRBBB 446 [quartiles 430-467] ms, iLBBB: 445 [quartiles 409-461] ms, cLBBB: 465 [quartiles 450-481] ms) were also longer in subjects with cRBBB, as well as in those with incomplete and complete LBBB (**Table 1**).

2D-echocardiographic assessment demonstrated significant, sex-independent structural differences in individuals with BBB, also after excluding subjects with prevalent diseases such as HF and MI. Compared to subjects with a normal electrical conduction, LVEF was significantly reduced in patients with cRBBB and even more pronounced in those with cLBBB (No BBB 58.4 [95% CI 55.0-61.3], cRBBB 57.3 [95% CI 54.9-60.7], cLBBB 54.5 [95% CI 51.0-57.1]). A significant reduction in right ventricular systolic function, assessed by fractional area change, was observed only in cRBBB. Chamber diameters showed a significantly increased interventricular septal thickness in both BBB types. Left ventricular end-diastolic diameter was enlarged in both BBB groups whereas right ventricular end-diastolic volume was particularly increased in cRBBB. Left atrial (LA) volumes were also increased in both BBB types, with the most pronounced enlargement observed in cLBBB. Additionally, in individuals with cLBBB we also observed a significantly lower LA-strain compared to the group without BBB (**Figure 3 and Table S2**).

**Figure 3:**
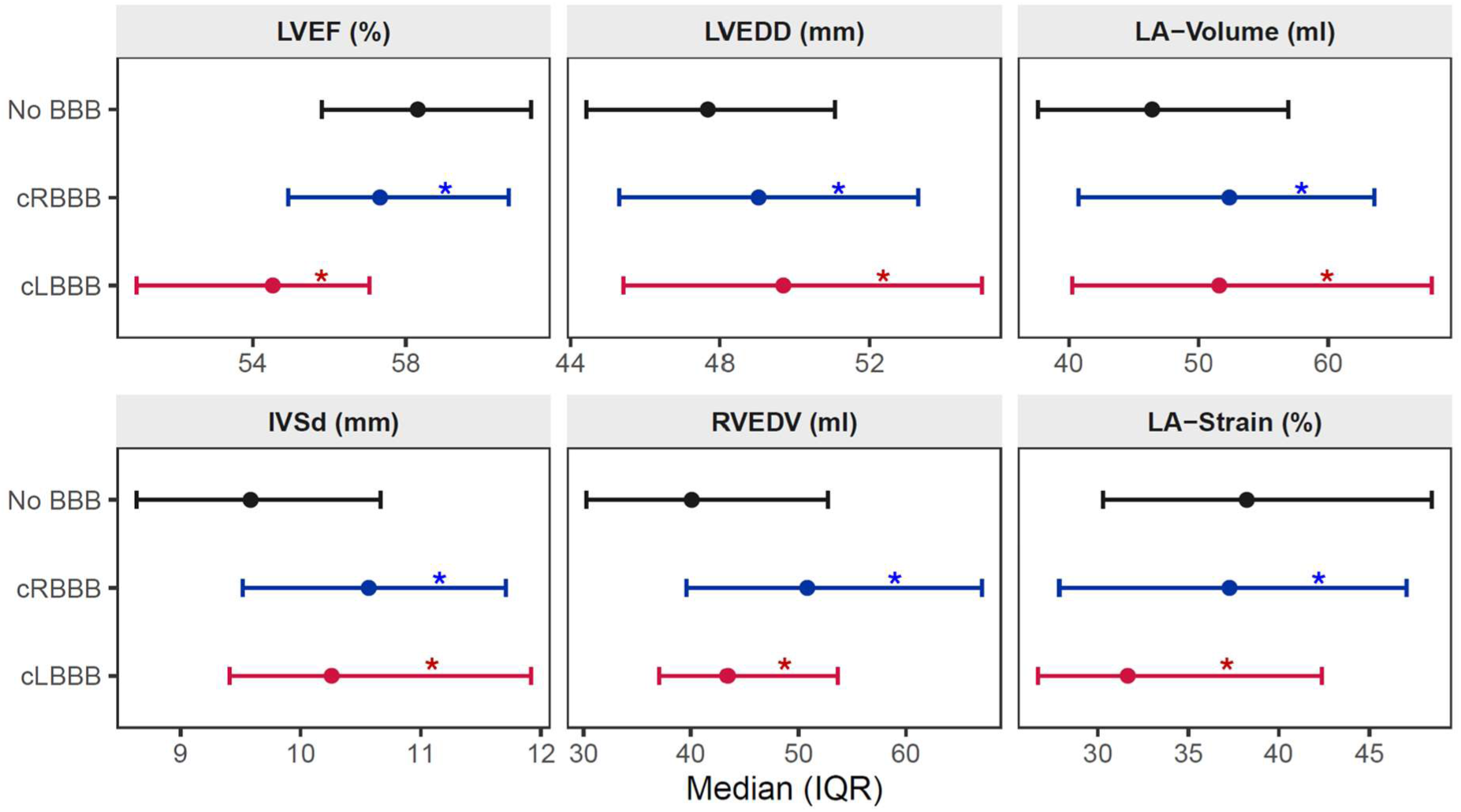
2D-Echocardiographic Parameters According to Bundle Branch Block Types. Forest plots displaying the median and quartiles (IQR) for key 2D-echocardiographic structural and functional parameters in individuals without BBB (black), as well as in subjects with RBBB (blue) and LBBB (red). The asterisks (*) indicate statistical significance compared with subjects without BBB, assessed via the Hodges-Lehmann method. *Abbreviations: cLBBB = complete left bundle branch block; cRBBB = complete right bundle branch block; IVSd = interventricular septal thickness at diastole; LA = left atrial; LVEDD = left ventricular end-diastolic diameter; LVEF = left ventricular ejection fraction; RVEDV = right ventricular end-diastolic volume*.

iRBBB demonstrated similar, albeit less pronounced, alterations in chamber diameters. Conversely, iLBBB showed more pronounced changes such as for example a median LVEF in the lower normal range (51.1 [95% CI 49.6-60.7]) (**Table S2**). Compared with individuals without conduction disturbances, the majority of patients with complete BBB (cRBBB, 77.7%; cLBBB, 78.3%) had at least one echocardiographic parameter outside the normal range.

### 3.5 Associations Between BBB and Long-Term Clinical Outcomes

In the total cohort, increasing QRS duration, treated as a continuous variable, measured in ms, was associated with a graded risk increase for the total cardiovascular outcomes. Each 10-ms increment in QRS duration above the median was associated with a 1.06-fold increase in risk (hazard ratio [HR], 1.06; 95% CI, 1.04-1.09; p < 0.001), with risk increasing even at QRS durations below the conventional ≥120-ms threshold for complete BBB (**Figure 4**).

**Figure 4:**
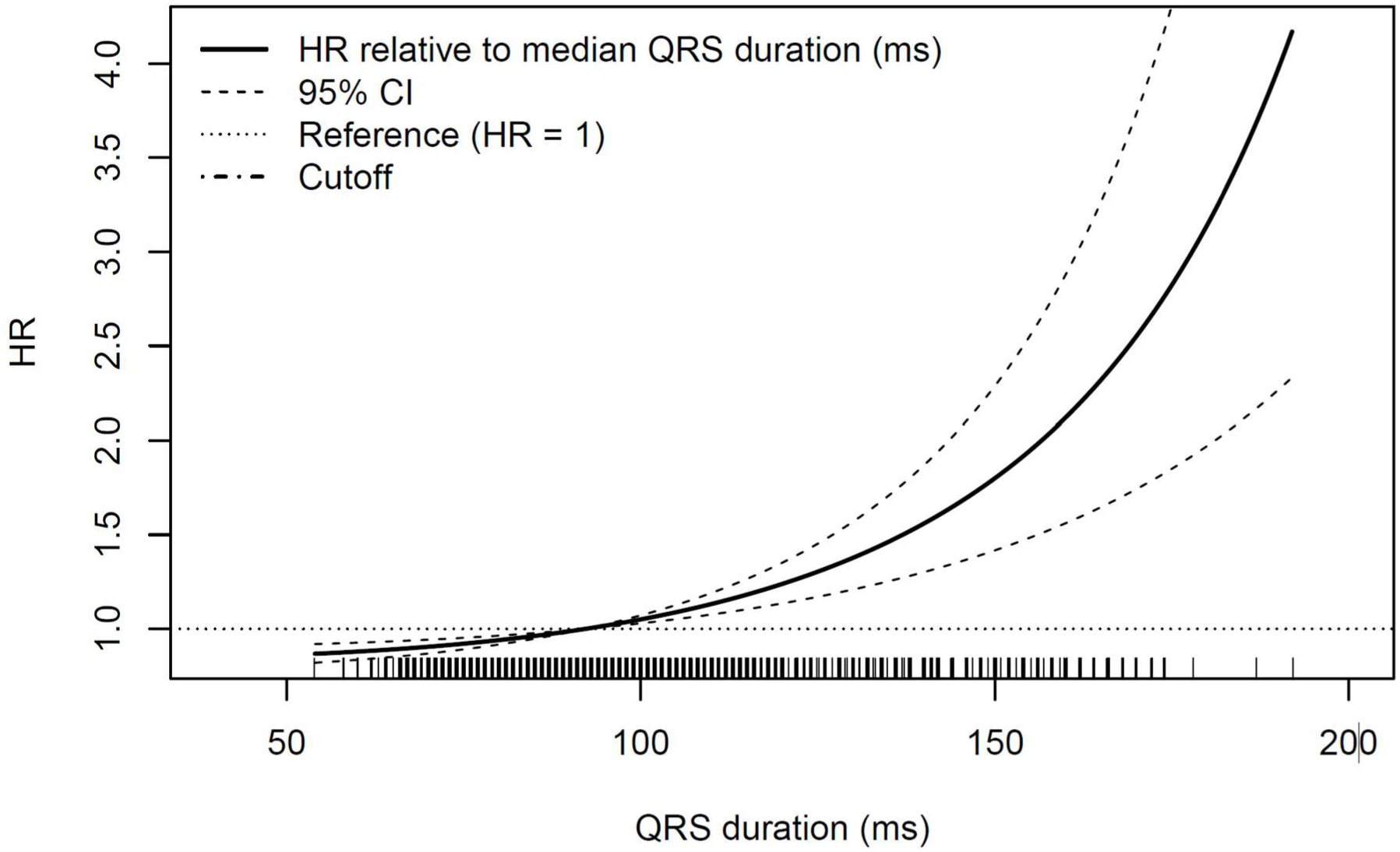
Continuous Association Between QRS Duration and the Total Cardiovascular Outcome. Total cardiovascular outcome: all-cause mortality, heart failure, atrial fibrillation, myocardial infarction, stroke, pulmonary embolism. Restricted cubic spline curve illustrating the estimated hazard ratio (solid line) as a continuous function of QRS duration. The dashed lines represent the 95 confidence intervals. The dotted horizontal line indicates the reference hazard ratio of 1.0, which is set at the median QRS duration. The tick marks on the x-axis (rug plot) depict the distribution of QRS duration values across the study population. *Abbreviations: CI = confidence interval; HR = hazard ratio; ms = milliseconds*.

During the follow-up of almost five years, both BBB types were associated with an increased risk of the total cardiovascular outcome compared with subjects without BBB, after adjusting for sex, and filtering out prevalent cardiovascular diseases (cLBBB: HR 2.71; 95% CI, 1.68–4.39; p <0.001; cRBBB: HR 1.52; 95% CI, 1.09– 2.12; p = 0.01). Individuals with cLBBB had a significantly increased risk of all-cause mortality as well, but no associations with the focused cardiovascular outcome (**Figure 5**). For both BBB types, the highest risks were observed for AF (cRBBB: HR 3.05 [95% CI 1.81 - 5.15], cLBBB: HR 4.15 [95% CI 1.95 - 8.84]), MI (cRBBB HR 2.78 [95% CI 1.31 - 5.93], cLBBB HR 5.48 [95% CI 2.25 - 13.35]) and HF (cRBBB HR 2.37 [95% CI 1.22 - 4.58], cLBBB HR 4.77 [95% CI 2.12 - 10.77]). Individuals with cRBBB had additionally exhibited a significantly higher risk of stroke (HR 2.8 [95% CI 1.44 - 5.46], **Figure 6**).

**Figure 5:**
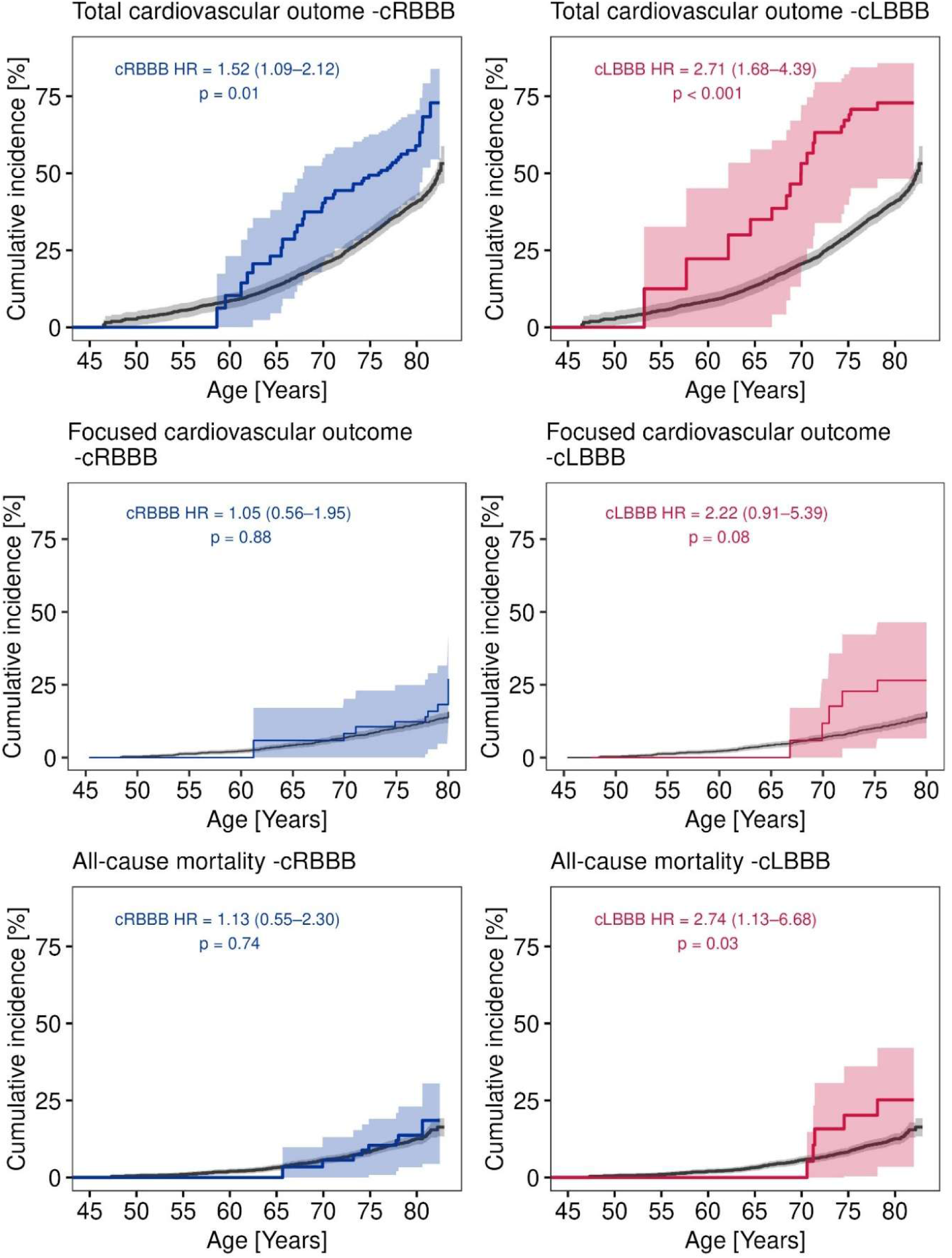
Cumulative Incidence of Clinical Outcomes According to Bundle Branch Block Subtypes. Cumulative incidence curves with age as the underlying time scale for (A+B) Total cardiovascular outcome (All-cause mortality, heart failure, atrial fibrillation, myocardial infarction, stroke, pulmonary embolism), (C+D) Focused cardiovascular outcome (cardiovascular mortality, myocardial infarction, stroke), (E+F) All-cause mortality. The solid lines represent the estimated cumulative incidence for subjects without a bundle branch block (black), with cRBBB (blue), and with cLBBB (red). The shaded areas indicate the corresponding 95% confidence intervals. *Abbreviations: HR = hazard ratio; cLBBB = complete left bundle branch block; cRBBB = complete right bundle branch block*.

**Figure 6:**
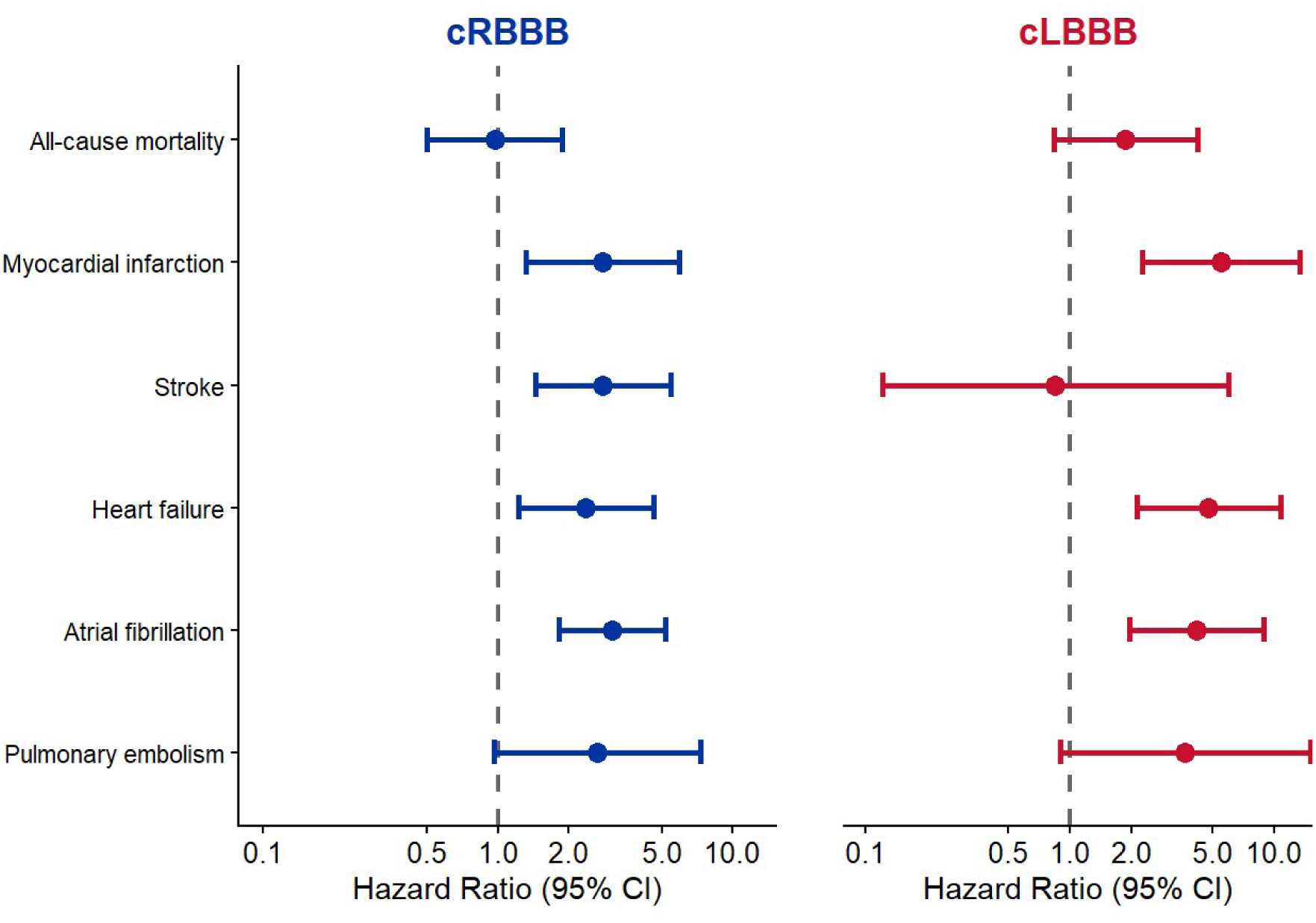
Hazard Ratios for Specific Clinical Outcomes Stratified by Bundle Branch Block Subtype. Forest plots displaying the risk of individual outcomes for individuals with RBBB (blue) and LBBB (red) versus those without bundle branch block. Data are presented as hazard ratios with 95% confidence intervals on a logarithmic scale. The dashed vertical line marks the reference value (hazard ratio = 1). *Abbreviations: CI = confidence intervals; cLBBB = complete left bundle branch block; cRBBB = complete right bundle branch block*.

## 4. Discussion

First, our study demonstrates that while RBBB exhibited a four-fold higher prevalence than LBBB in this cohort study of the general population of Hamburg, both conduction disturbances were associated with a substantial cardiovascular risk burden. Second, individuals with either BBB subtype demonstrated a higher prevalence of traditional cardiovascular risk factors, coronary artery disease, and HF. Third, the presence of a BBB correlated with structural echocardiographic abnormalities. cLBBB was specifically associated with chamber enlargements and reduced ejection fraction. Fourth, a longer QRS width was associated with a worse prognosis, even at values below 120ms. Fifth, both cRBBB and cLBBB conferred an increased risk of the total cardiovascular outcome. Compared with cRBBB, cLBBB was associated with a 50% higher overall risk of adverse events and an approximately twofold higher risk of all-cause mortality.

### 4.1 Subclinical Remodeling and Prognostic Implications of BBB

To our knowledge, this is the first study in a general population cohort to concurrently evaluate the prognostic significance of BBB and characterize associated subclinical structural alterations using comprehensive echocardiographic phenotyping. In this middle-aged cohort, the prevalence of BBB was relatively low, which aligns with previous observations indicating that BBB prevalence increases primarily after 80 years of age.^13^ We observed a markedly higher comorbidity burden among individuals with BBB, excluding iRBBB. Given this elevated risk profile, the presence of BBBs could be a valuable screening tool in primary care. Particularly in a middle-aged population, early identification provides a critical window of opportunity to implement targeted interventions and optimize the cardiovascular risk profiles in affected patients. While prior investigations have associated cLBBB and cRBBB to diabetes mellitus and hypertension, our data extend these findings by additionally demonstrating an association with hypercholesterolemia, a risk factor not explicitly evaluated in earlier BBB cohorts.^15,16^

A central novelty of our analysis is the systematic echocardiographic assessment, which revealed significant structural and functional alterations in subjects with BBBs compared to the total cohort. Although absolute values largely remained within normal physiological limits, LVEF was lower in both BBB morphologies, with a more pronounced reduction observed in LBBB. Furthermore, we noted subclinical divergences in ventricular diameters and atrial dimensions in most of the complete BBBs. Importantly, these structural alterations persisted even after the exclusion of individuals with prevalent cardiovascular diseases. Historically, echocardiographic data pertaining to BBB have predominantly been derived from older registry studies or subgroup analyses, which primarily established the association between LBBB, reduced LVEF, and altered LV diameters.^4,17^ Building upon this foundation, our findings newly demonstrate that cRBBB is similarly associated with lower LVEF, and that BBBs show subclinical atrial and ventricular remodeling.

Our finding on long-term outcomes corroborate data from an older Finnish cohort of middle-aged subjects, confirming that a progressive increase in QRS duration is significantly associated with higher mortality risk.^18^ For cLBBB, the existing literature predominantly highlights established associations with cardiovascular mortality and incident HF.^8,19^ Our analysis expands upon this prognostic profile by demonstrating an association with all-cause mortality as well and identifying cLBBB as an independent predictor of incident MI. Conversely, while recent literature has associated RBBB with all-cause mortality and an increased risk of pacemaker implantation, our longitudinal data demonstrated a significantly increased risk of incident stroke with cRBBB within five years of follow-up.^9^

### 4.2 Clinical Implications

Our observation of structural alterations even in individuals without prevalent cardiovascular disease suggests a complex, potentially bidirectional relationship between conduction disturbances and myocardial function. On one hand, BBB may represent an early subclinical manifestation of incipient HF, a hypothesis supported by our longitudinal outcome analyses.^20^ On the other hand, it is equally plausible that the conduction delay acts as a primary driver of myocardial remodeling. The resulting electrical dyssynchrony and subsequent mechanical phenomena, such as “septal flash” and “apical rocking”, may act as potential precursors to adverse structural remodeling.^4^ This mechanical theory is further reinforced by the profound clinical response to cardiac resynchronization therapy observed in patients with cLBBB, where restoring synchrony often reverses these remodeling processes.^10^

From a clinical perspective, the incidental detection of a complete LBBB or RBBB, excluding iRBBB, should prompt intensive screening and optimization of cardiovascular risk factors. Given that subclinical structural changes were evident even in asymptomatic individuals, our findings suggest that a baseline echocardiographic assessment could be considered to identify those at the highest risk for progression, for example to overt HF. Particularly in middle-aged populations, such a proactive approach may provide an opportunity for early intervention before potentially irreversible structural changes occur. The incidence of cardiovascular events was particularly high among patients aged ≥80 years in our study. This raises the possibility that changes remaining subclinical earlier in life might become more pronounced with advancing age, potentially making complete BBB an early sensitive marker for the later development of structural heart diseases.

Beyond ventricular alterations, we observed significant atrial structural remodeling and a heightened risk of incident AF across both BBB morphologies. cRBBB was uniquely associated with an increased risk of incident stroke. This finding highlights the potential clinical value of intensified AF screening in patients with cRBBB, using tools such as smartwatches or prolonged Holter monitoring.^21^ Identifying subclinical AF early in this population would offer a crucial opportunity to initiate timely anticoagulation and mitigate stroke risk.

### 4.3 Limitations

Several limitations warrant consideration. First, the observational design precludes definitive causal inference, and residual confounding may persist despite multivariable adjustment. Second, BBB status was assessed at a single baseline timepoint using resting 12-lead ECGs. Consequently, incident conduction abnormalities or temporal changes during the follow-up period could not be evaluated. Third, the relatively small number of subjects with LBBB, particularly iLBBB, limited statistical power for certain subgroup analyses and may lead to wider CIs. Importantly, this low absolute number accurately reflects real-world epidemiologic distributions, reinforcing the applicability of our findings to routine screening scenarios in the general population. Fourth, outcome ascertainment relied in part on self-reported data and registry linkages, which may introduce misclassification bias, despite careful adjudication of key endpoints. Fifth, while the relatively short follow-up period may have limited the evaluation of very long-term trajectories, it is notable that significantly elevated risks of adverse cardiovascular outcomes among individuals with BBB were already robustly detectable within this limited timeframe. Finally, the cohort comprised middle-aged adults from a single metropolitan region, which may limit generalizability to other geographic, ethnic, or older populations. Given the established age-dependent increase in both structural heart disease and conduction disturbances, it is highly plausible that the structural deviations and prognostic implications of BBB would be even more pronounced in an older population.

## 5. Conclusion

In this large population-based study focusing on ECG conduction abnormalities, longer QRS duration was associated with poorer cardiovascular outcomes at long-term follow-up. Both complete cLBBB and cRBBB were associated with a higher prevalence of cardiovascular risk factors as well as structural cardiac changes, as reflected in 2D-echocardiographic parameters of either right, left ventricular or both systolic function and chamber sizes. Importantly, even after adjustment for age, sex, and prevalent cardiovascular diseases, cLBBB and cRBBB remained associated with increased risks of the total cardiovascular outcome. These findings indicate that the presence of a BBB identifies individuals in the general population at higher cardiovascular risk, suggesting the need for further diagnostic work-up or closer follow-up to improve long-term outcomes.

## Data Availability

The data underlying this article are available in the article and in its online supplementary material.

## Acknowledgements

The authors acknowledge the subjects of the HCHS and cooperation partners, patrons and the Deanery from the University Medical Centre Hamburg - Eppendorf for supporting the HCHS. Special thanks apply to the staff at the Epidemiological Study Centre for conducting the study. The participating institutes and departments from the University Medical Center Hamburg-Eppendorf contribute all with individual and scaled budgets to the overall funding of the HCHS. The HCHS is additionally funded by the Kühne Foundation, the Max Delbrück Center - Berlin and the Innovative medicine initiative [Grant Number 116074]. The HCHS is further supported by „Deutsches Zentrum für Herz-Kreislauf-Forschung (DZHK)“; „Deutsche Stiftung für Herzforschung“; Joachim Herz Stiftung“; „Seefried Stiftung” Siemens (Healthineers AG); by the authorities of the City of Hamburg and by donations from the “Förderverein zur Förderung der HCHS e.V.”. Sponsor funding has in no way influenced the content or management of this study.

## Sources of Funding

Although no dedicated funding was received for this specific analysis, the overarching HCHS is financed through individual and scaled contributions from the participating institutes and departments of the University Medical Center Hamburg-Eppendorf. The HCHS also receives financial support from the euCanSHare grant agreement [Grant Number 825903-euCanSHare H2020], the Joachim Herz Foundation, the Foundation Leducq [Grant Number 16 CVD 03], the Kühne Foundation, and the Innovative Medicines Initiative [Grant Number 116074]. Further backing is supplied by the “Deutsche Gesetzliche Unfallversicherung (DGUV)”, “Deutsches Krebsforschungszentrum (DKFZ)”, “Deutsches Zentrum für Herz-Kreislauf-Forschung (DZHK)”, “Deutsche Stiftung für Herzforschung”, “Seefried Stiftung”, Bayer, Amgen, Novartis, Schiller, Siemens, Topcon, Unilever, TePe® (2014), alongside charitable donations from the “Förderverein zur Förderung der HCHS e.V.”. The financial sponsors played no role in the design, execution or management of the study.

## Conflict of Interest

Juliana Senftinger declares no conflicts of interest. Vivian Link declares no conflicts of interest. Matthias Klimek declares no conflicts of interest. Laura Dorothea Keil declares no conflicts of interest. Maria Luisa Benesch Vidal declares no conflicts of interest. Jan Rieß declares received speaker honoraria from AstraZeneca. Raphael Twerenbold reports research support from the German Center for Cardiovascular Research (DZHK), the Kühne Foundation, the Joachim Herz Foundation, the Swiss National Science Foundation (Grant No P300PB_167803) and the Swiss Heart Foundation as well as speaker honoraria/consulting honoraria from Abbott, Amgen, Astra Zeneca, Daiichi-Sankyo, Psyros, Roche, Siemens, Singulex, Spinchip and Thermo Scientific BRAHMS, all outside the submitted work. Raphael Twerenbold is co-founder and shareholder of the ART-EMIS Hamburg GmbH, which holds an international patent application on a computing device for estimating the probability of *MI* (International Publication Numbers WO2022043229A1, TW202219980A).

Christina Magnussen received funding from the German Center for Cardiovascular Research (DZHK) and the Deutsche Stiftung für Herzforschung unrelated to the current work. CM received speaker fees from Edwards, AstraZeneca, Novartis, Boehringer Ingelheim/Lilly, Bayer, and Novo Nordisk outside this work. CM has participated in advisory boards for Boehringer Ingelheim, Alnylam and Novo Nordisk. Renate B. Schnabel declares no conflicts of interest. Stefan Blankenberg declares no conflicts of interest. Andreas Ziegler declares no conflicts of interest. Peter Moritz Becher reports reports speaker fees from AstraZeneca and Ingelheim Boehringer, all outside the submitted work. P.M.B. received funding from the German Research Foundation, outside the submitted work. Peter Clemmensen reports consultant fees from Acarix AB, is an investigator for Acarix AB and Bayer Healthcare, and is on End Point Review Committees for Boehringer Ingelheim and WGC, outside the submitted work.

## Abbreviations

AF: Atrial fibrillation
AUC: Area under the receiver operating characteristic curve
BBB: Bundle branch block
CI: Confidence interval
cLBBB: Complete left bundle branch block
cRBBB: Complete right bundle branch block
ECG: Electrocardiogram/Electrocardiographic
HCHS: Hamburg City Health Study
HF: Heart failure
HR: Hazard ratio
iLBBB: Incomplete left bundle branch block
iRBBB: Incomplete right bundle branch block
LA: Left atrial/atrium
LVEF: Left ventricular ejection fraction
MI: Myocardial infarction
ms: Milliseconds
SD: Standard deviation
TTE: Transthoracic echocardiography
2D: 2-Dimensional

## Illustrations and Figures

The authors do hereby declare that all illustrations and figures in the manuscript are entirely original and do not require reprint permission.

## Notes

### Clinical Trial

NCT03934957

### Author Declarations

The ethics committee of the State of Hamburg (Chamber of Medical Practitioners, reference number PV5131) did not object against the conduct of the study, and the respective Data Protection Commissioners agreed to the participation of the study. All subjects provided written informed consent.

